# Physician Perceptions of Surveillance: Wearables, Apps, and Bots for COVID-19

**DOI:** 10.1101/2021.04.26.21256102

**Authors:** Alexandra R. Linares, Katrina A. Bramstedt, Mohan M. Chilukuri, P. Murali Doraiswamy

## Abstract

**Objective:** To characterize the global physician community’s opinions on the use of digital tools for COVID-19 public health surveillance and self-surveillance.

**Methods:** Cross-sectional, random, stratified survey done on Sermo, a physician networking platform, between September 9-15, 2020. We aimed to sample 1,000 physicians divided among the USA, EU, and rest of the world. The survey questioned physicians on the risk-benefit ratio of digital tools, as well as matters of data privacy and trust.

**Results:** The survey was completed by 1004 physicians with a mean (SD) age of 49.14 (12) years. Enthusiasm was highest for self-monitoring smart watches (66%) and contact tracing apps (66%) and slightly lower (48-56%) for other tools. Trust was highest for health providers (68%) and lowest for technology companies (30%). Most respondents (69.8%) felt that loosening privacy standards to fight the pandemic would lead to misuse of privacy in the future.

**Conclusion:** The survey provides foundational insights about how physicians think of surveillance. Collaborations between public health and technology researchers to strengthen evidence of effectiveness and build public trust may be useful.

## Introduction

Public health surveillance is the systematic collection and analysis of health-related data to prevent or control disease, followed by its application for public health action.^1^ The global scale of the COVID-19 pandemic has accelerated the use of non-traditional, technology-based, public health, and self-surveillance mechanisms to control the spread of SARS-CoV-2.^2-23^ Examples of such tools include contact-tracing apps, analyses of global positioning systems and social media data for population movement tracking, fever-sensing infrared thermal detection systems (ITDS), symptom self-screeners (e.g., chatbots), and smart watch applications to detect physiological signs of infection.^4-8,10^

Digital technologies have the ability to rapidly collect, store, analyze, and share numerically encoded information, making them potentially highly useful in a pandemic such as COVID-19. Blue Dot, a Canadian digital health company, reportedly identified the emergence of COVID-19 through aggregation of big data from sources such as social media and air travel, before even the WHO issued an alert.^14^ However, these digital surveillance tools are experimental, and their accuracy across different settings is not fully established.^15-20^ For example, studies have shown that the accuracy of facial recognition technologies differs by race, gender and age.^22^ These tools also come with a number of potential legal and ethical risks,^18,24-28^ such as privacy concerns, discrimination, and over-reach of the data mission that “highlight the long-standing tensions between individual and collective rights”.^18^

Notably, the morbidity and mortality of the COVID-19 pandemic has heightened worldwide anxiety to an extent that digital public health surveillance has become ubiquitous (e.g., national requirements for downloading contact tracing apps; thermal scanning by employers and private businesses; personal location data collection via QR codes; texting of COVID-19 assay results to patients). COVID-19 is not the world’s first pandemic, nor will it be the last. Thus, it is vital to understand the views of physicians, as they are involved in many facets of health data and its application to COVID-19 care. The aim of this report is to characterize the views of physicians regarding the benefits and risks of surveillance technologies.

## Methods

### Study sample

To characterize the opinions of physicians on this topic, we analyzed data from a cross-sectional, random, stratified survey of physicians registered with Sermo, a secure digital platform for medical crowdsourcing and anonymous surveys. The Sermo platform is exclusive to verified and licensed physicians and has over 800,000 registered physicians, of all specialties, worldwide.

Following informed consent, the English-language survey sampled physicians between September 9 and September 15, 2020 (prior to initiation of SARS-CoV-2 vaccination), with a target sample size of 1000 doctors equally divided between the US, EU, and rest of the world (RoW). The survey results were de-identified to create anonymized data for analyses. This was deemed as exempt research by Duke University Medical Center’s Institutional Review Board.

### Survey instrument

Five questions in the survey (Figure 1, Supplemental Table 1) asked physicians their opinions on the benefits and risks/harms of using smart watch sensor alerts, contact tracing apps, thermal cameras for mass fever screening, chatbots, and social media tracking for public health or self-surveillance. These questions focused on risk-benefit ratio, and the answer options were “Yes”, “No”, and “Uncertain”. For the questions on sensors (wearables and thermal cameras), we provided accuracy estimates derived from published studies. We specified that informational chatbots do not require regulatory approval in most countries for the question on chatbots, as physicians may not be aware of this. The survey then asked about level of trust in different organizations (technology companies, government, employer, medical provider, educational universities/non-profit bodies, or no one) to protect private surveillance data. The answer choices for this question were “Very much”, “Somewhat”, “Neutral”, “Not really”, or “Not at all”. Respondents were then asked about the impact of current surveillance on future privacy standards. A final question asked physicians to provide brief qualitative comments to elaborate on their views. The results of two survey questions are reported elsewhere.^29,30^

**Figure 1.**
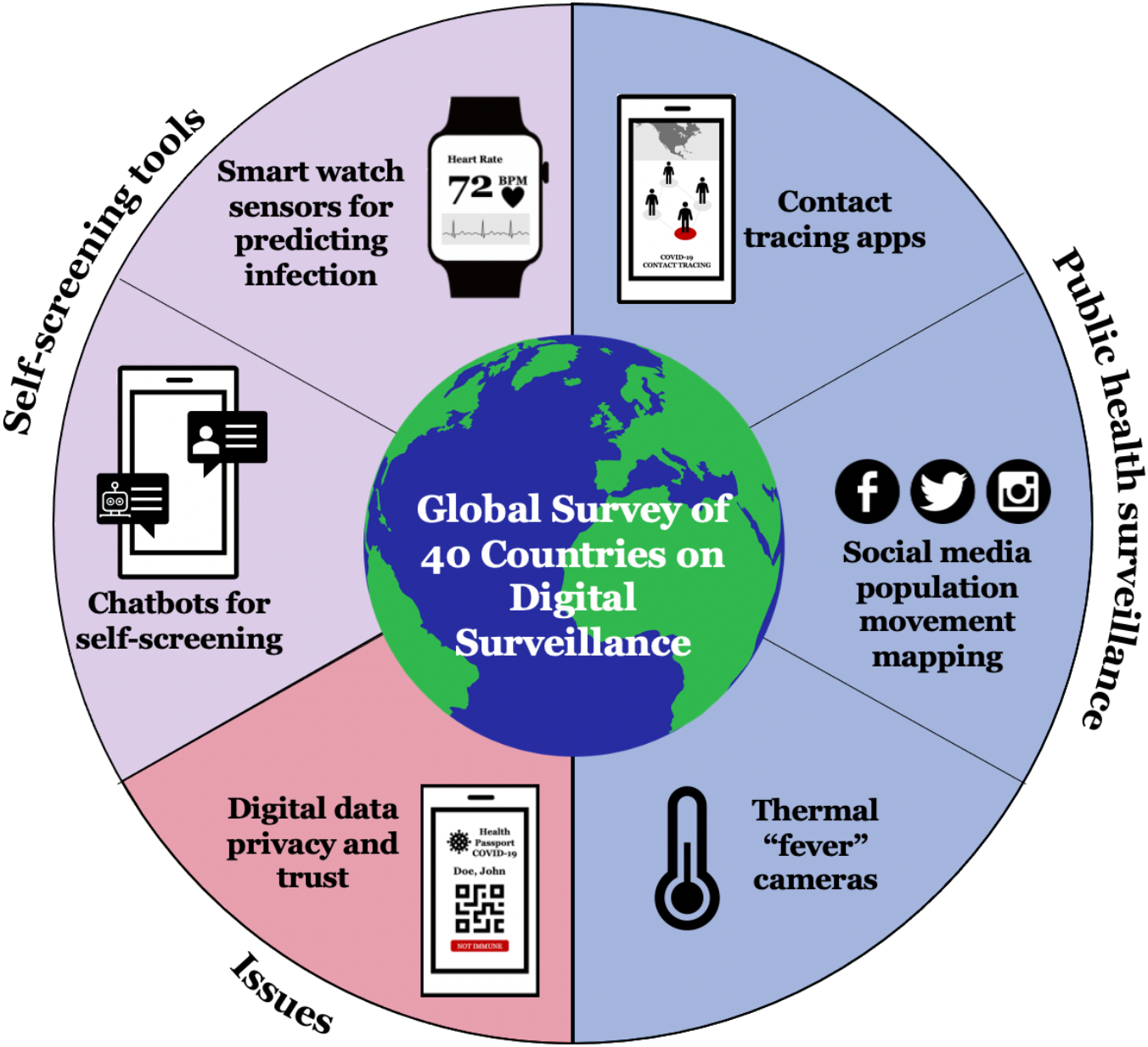
Schematic illustration of surveillance tools and issues queried in the survey. The survey examined the risk-benefit ratio of two self-screening (purple) and three public health (blue) surveillance digital tools. It also addressed issues around trust and misuse (red).

### Data and statistical methods

Descriptive statistics examined physicians’ characteristics and opinions by age group, gender, frontline status, and geographic region. To test the effect of age, subjects were grouped as “younger” or “older” by age 49 years. To test the effect of frontline status, physicians more directly involved in COVID-19 care were grouped as frontline (e.g., internal medicine, ICU, ED), whereas the rest were categorized as non-frontline (although we recognize that all physicians may interact with or consult on COVID-19 patients). For geographic analyses, we pooled doctors into three groups based on location of practice (US, European Union, rest of the world), while recognizing these subgroups are not homogenous. The five categories relating to trust were combined into three categories as trusted (“Somewhat” or “Very much” responses), not trusted (“Not really” or “Not at all”), and “Neutral”. Gender analyses were restricted to those who categorized themselves as male or female. ANOVA, t-test, and Chi-square tests with *P* values <.05 were viewed as qualitatively different. As this was an exploratory study, we did not adjust for small cell sizes or multiplicity. We used JMP Pro 15 (SAS), as well as Protobi.

## Results

### Sample characteristics

The final respondent sample consisted of 1004 physicians representing 40 countries in North and South America, Europe, and Asia-Pacific (Supplemental Table 2). The average age of the sample was 49.1 ± 12.3 years, and 49% of respondent physicians were characterized as frontline. Of the sample, 40% were male, 20.6% were female, and 39% opted out of indicating their gender.

### Utility of Smart Watches for Infection Detection

The majority of respondents (65.5%) felt that the possible benefits of infection detection alerts from smart watch sensors would outweigh the possible risks/harms (Figure 2), 17.3% felt that the possible benefits would not outweigh the possible risks (*X*^2^ = 468.58, *p*<.0001), and another 17.1% were uncertain. Younger physicians (69.3%) were more likely to support the use of smart watch sensors compared with older physicians (61.3%) (*X*^2^=7.06, *P*=.03). Responses did not differ significantly between males and females, or by frontline status.

**Figure 2.**
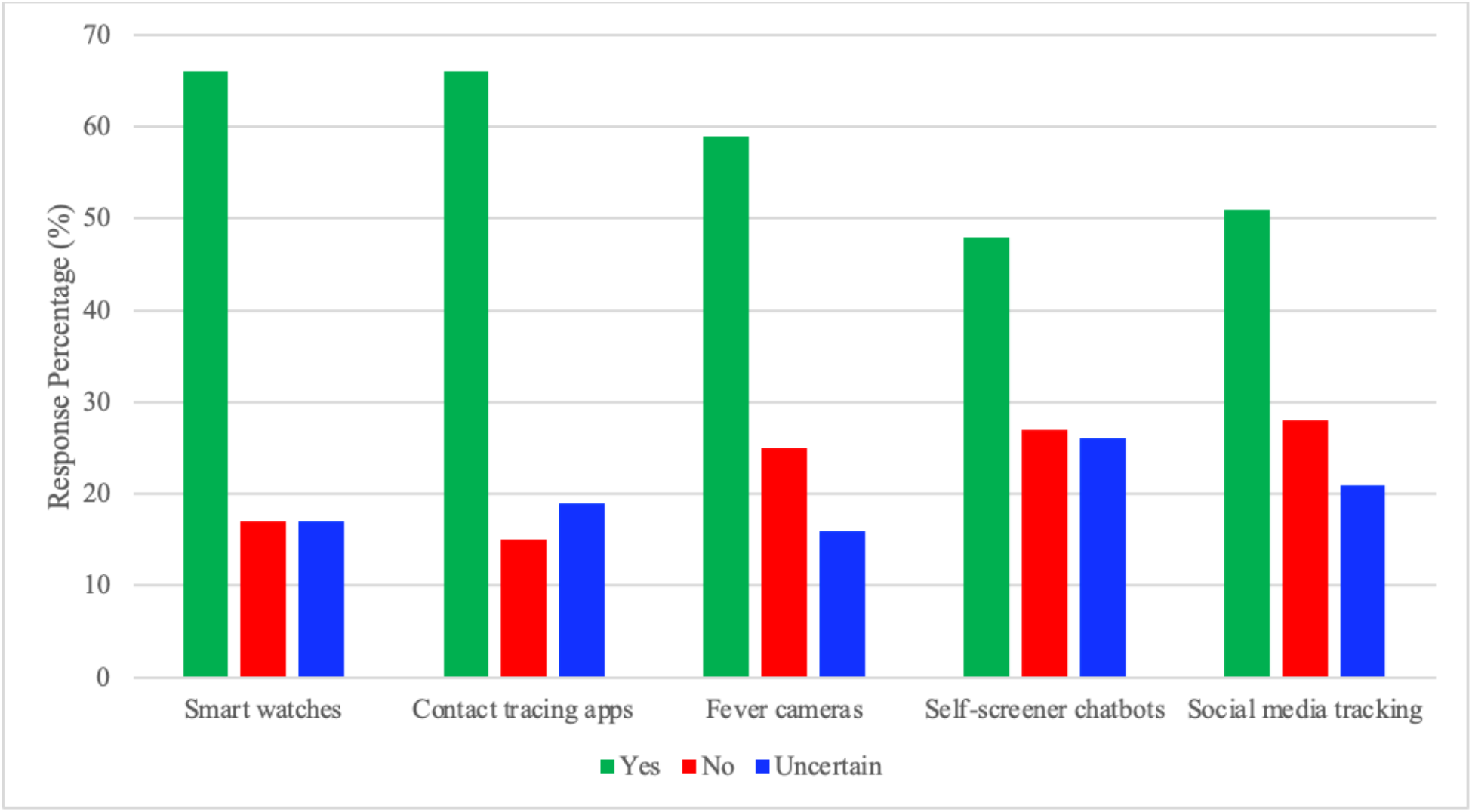
Physician Perceptions of Digital Surveillance Tools relevant to COVID-19. Graph illustrates percent of respondents who were supportive (green bars), uncertain (blue bars) or unsupportive of the use of surveillance tools. Please see text for statistical differences.

### Utility of Contact Tracing Apps

Most respondents (66.4%) felt that the possible benefits of contact tracing apps would outweigh the possible risks (*X*^2^ = 496.85, *P* < .0001) (Figure 2), with 15.0% responding that the benefits would not outweigh the risks and 18.5% uncertain. Younger physicians (69.7%) were marginally more likely to support it compared to older ones (62.8%) (*P=*.02). Responses did not differ significantly by frontline status.

### Utility of Infrared Thermal “Fever Cameras”

Just over half of respondents (58.9%) felt that the benefits of mass fever screenings would outweigh the possible risks (*X*^2^ =306.34, *P* < .0001). A quarter of respondents (25.0%) said the potential benefits would not outweigh the possible risks, and 16.1% were uncertain (Figure 2). Younger physicians (63.6%) were more likely to support the use of mass fever screenings compared with older physicians (53.6%) (*X*^2^ = 11.62, *P*=.003), whereas older physicians were more likely to be uncertain. Male physicians (60%) were slightly more likely to support than female physicians (51%) (*P*=0.012). Responses did not differ by frontline status.

### Utility of Symptom Screener Chatbots

Less than half of respondents (47.6%) felt that the benefits of symptom self-screener chatbots would outweigh the possible risks (*X*^2^ = 92.18, *P*< .0001). The other half of respondents were split, with 26.6% feeling that the potential benefits would not outweigh the possible risks and 25.8% uncertain (Figure 2). Responses did not significantly differ between males and females, nor by physician age or frontline status.

### Utility of Social Media for Population Movement Tracking

About half of respondents (50.9%) felt that the benefits of using social media postings to create COVID-19 symptom maps and population movement maps would outweigh the possible risks, (*X*^2^ = 145.68, *P* < .0001) whereas 27.8% felt the potential benefits would not outweigh the possible risks, and 21.3% were uncertain (Figure 2). Younger physicians (56.1%) were more likely to support the use versus older physicians (45.2%) (*X*^2^ = 14.34, *P*=.0007), and older physicians tended to be more uncertain. Responses did not differ by gender or frontline status.

### Which Entity Do You Trust the Most with Your Personal Surveillance Data?

Physicians picked “medical providers” as the most trusted entity to protect privacy of COVID-19 surveillance data, with about 68% of respondents reporting that they trusted their medical provider (Figure 3). The second most trusted group was “educational/non-profit bodies”, with a combined 52% of respondents reporting “somewhat” and “very much” levels of trust. Conversely, the most distrusted group was “technology companies”, with only 30% of respondents reporting “somewhat” or “very much”, and 46% reporting “not really” or “not at all”. Following technology companies, respondents reported low levels of trust for the “government”, with only 36% responding “somewhat” or “very much”. Older physicians were more likely to be distrustful of technology companies (48.9%), the government (44.5%), and educational universities/non-profit bodies (26.9%) compared with younger physicians (42.4%, 33.9%, 16.9% respectively) (*P*=.038, .001, <.001). US physicians (54.1%) were more likely to be distrustful of technology companies, compared with both EU (41.0%) and RoW (41.3%) physicians (*p*<.001). US physicians (51.8%) were also more distrustful of the government, compared with RoW (38.6%) and EU (27.5%) physicians (*P*<.001).

**Figure 3.**
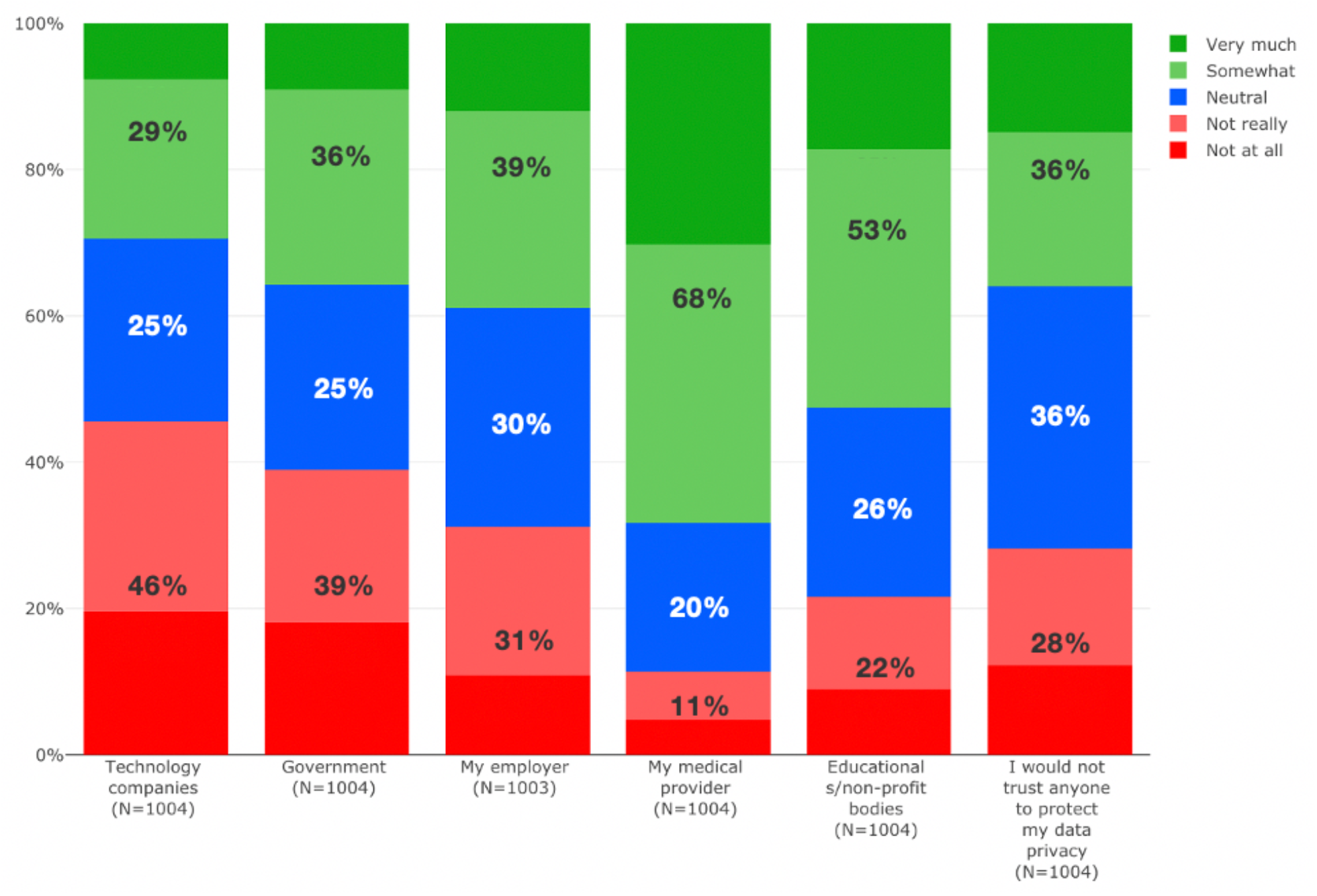
Physicians Perceptions of Trust in Various Entities to Protect their Personal Data. Graph illustrates that trust was highest for medical providers and lowest for tech companies. The colors show the actual percentages reported by each entity for the 5 level of trust categories. The percentages reported inside the bars combine the “Somewhat”/“Very much” and “Not really”/“Not at all” categories.

### Effect of Current Surveillance on Future Misuse of Privacy

The majority of respondents (69.8%) believed that potentially loosening privacy standards to fight the pandemic would lead to misuse of privacy in the future (*X*^2^ = 601.50, *P*< .0001) (Figure 4). Frontline physicians (73.2%) were more likely to voice concern compared with non-frontline physicians (66.5%) (*X*^2^ = 7.65 *P*=.022). More male physicians (71.9%) believed that a loosening of privacy standards would lead to misuse, compared with female physicians (61.8%) (*X*^2^ = 9.58 *P*=.048).

**Figure 4.**
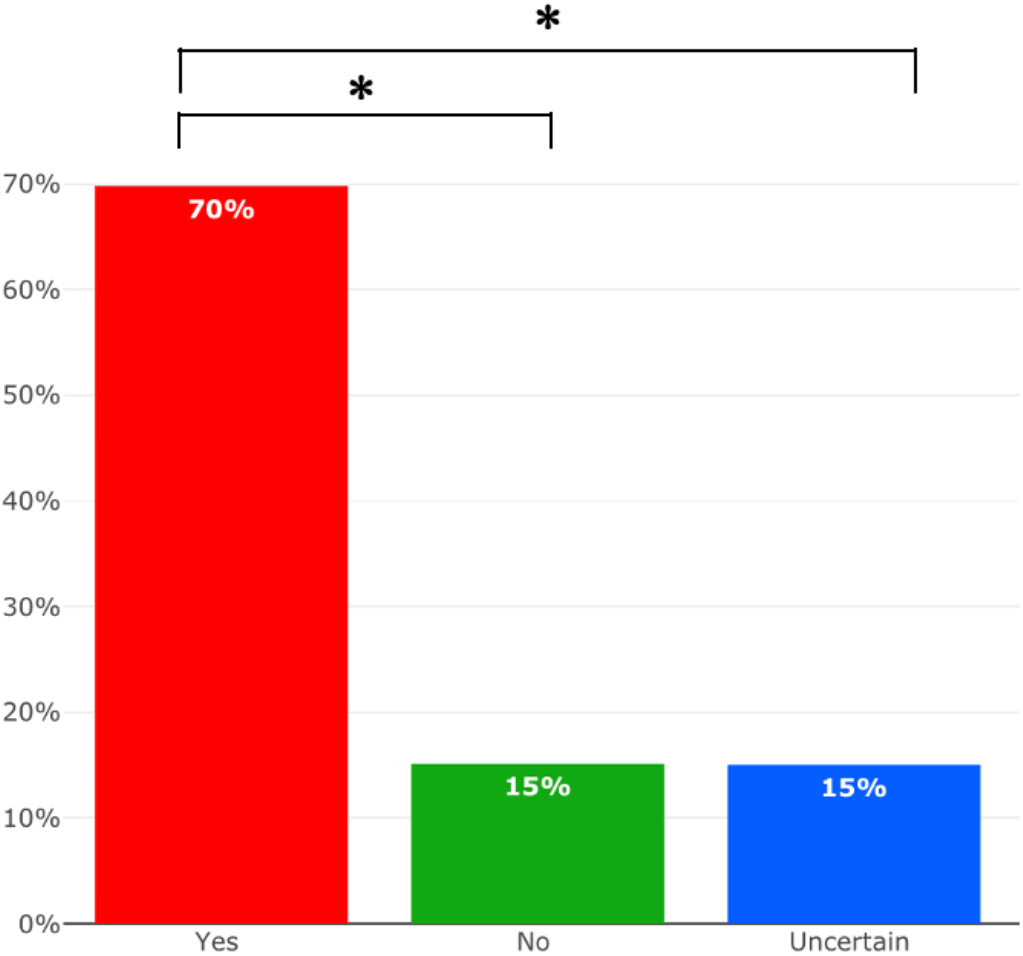
Physician Perceptions of the Risk for Future Misuse of Surveillance Data. Red bar illustrates the percent of respondents who *agreed* that loosening of privacy laws to fight the current pandemic would result in future misuse of personal data. Green bar indicates those who disagreed and blue bar indicates those who were uncertain. * indicates *P*<.05

### Selected Qualitative Comments by Physicians About Digital Surveillance

Respondents also had an option to provide qualitative comments on digital surveillance. Supportive comments (Supplemental Table 3) included statements such as “*the Future is here”*, “*must be made mandatory”*, “*anything that prevents deaths is fine, I don’t worry about privacy”*, and “*During the 1940-41 bombing of London called The Blitz, I believe there were zero residents of London who said ‘I have a constitutional right to leave my lights on a night if I feel like it*.’”. Concerns about efficacy (Supplemental Table 4) included statements like “*way too early”*, “*bad for patient and physician”*, and “*data should be analyzed in clinical trials”*. Concerns about harms (Supplemental Table 5) included statements such as “*Pandora’s box”*, “*creepy, extreme slippery slope”*, “*any great idea can have unforeseen* consequences” and “*I fear the behavior of people not technologies”*.

## Discussion

Data is currency. Technology companies know this, governments know this, and so does the public. Like with any currency, data can be accumulated, bought and sold, or even be stolen. Hence, its storage needs to be secure. Health data is a form of personal information that generally people want to keep private, and many regulations have been implemented to safeguard personal data privacy rights.^31,32^ During a pandemic, public health interests allow for broader powers, governments, and health systems to collect, use, store, and share personal information. However, as our survey shows, this creates concern among even the physicians who are part of this process (and concurrently attempting to prevent and treat the implicated illness).

Overall, support varied from 48% to 66% for the various surveillance tools. Two-thirds of physicians voiced support for the use of smart watches in self-monitoring. This appears consistent with recent studies documenting the promise of consumer smart watch based physiological signals (e.g., heart rate, sleep, activity, skin temperature) for discriminating COVID-19 test positive cases from negative cases, as well as for detecting pre-symptomatic COVID-19 infection.^4,5^ Further, a smartwatch linked platform, Aura, recently received an EU CE mark for this purpose based on its sensitivity of 94% and ability to detect an infection signal on average 2.64 days after inoculation.^6^ The minority of respondents who oppose smart watch based infection detection technology were likely concerned about the potential for noisy data leading to misdiagnosis and unnecessary testing.^19^

Two-thirds of physicians also voiced support for contact tracing apps, even those that collected personal data. Many countries have implemented contact tracing apps, and physicians are well versed in traditional contact tracing principles for infection control, both of which likely increased physician confidence in their utility. However, since our survey, some studies have questioned the effectiveness (e.g., sensitivity of only 7% in one study) and ethics of digital contact tracing.^20-22^ This suggests that the optimism of respondents in our survey may have been premature.

Physician support was slightly lower (59%) for “fever cameras” but still optimistic, consistent with the utility for mass screening offered by their high negative predictive value,^8^ However, the positive predictive value (less than 20% in one study) of these systems remains low,^8,9^ suggesting the need for further optimization to reduce false positives.

Support for the use of social media tracking (51%) and chatbots (48%) was also slightly lower. Social media tracking is a promising tool that offers real time data for public health officials to monitor citizen movement or social interactions during lockdowns.^10-13^ However, questions remain about lack of consent, accuracy, and misuse potential. Chatbots, especially those designed using WHO or CDC guidelines, very likely helped large numbers of users (over 200 million messages by some estimates) quickly get reliable information. However, to our knowledge, there are no published accuracy or outcomes data on the utility of chatbots for pandemic self-screening. Hence, there is a need for further research into effectiveness and potential for spread of misinformation.^20,21^

Respondents in our survey also voiced concerns over privacy risks and over-reach of the data mission. Respondents had low trust in technology companies (30%) and governments (36%) to safeguard surveillance data. Trust was highest with medical providers (68%), followed by non-profit organizations. The higher level of trust among EU physicians may be due to the stricter data privacy laws in the EU versus the US.^31^ These concerns are legitimate, since some technology platforms rely on selling user data to advertisers,^27^ and studies have found apps and chatbots share information with a variety of third parties.^20,27-29^ The risks also go beyond privacy breaches.^27^ Historically, surveillance has worsened stigmatization and discrimination against racial or religious minorities who were often falsely blamed for disease outbreaks.^27^ Further, some governments have reportedly used the pandemic to rank citizens by health status or analyze personal telecommunications traffic.^27^ Hence, surveillance done wrong may “invite mission creep into adjacent fields, such as automated policing and content control”.^24^

“No turning back” is a famous quote used in many settings, and our research makes it pertinent to digital health as well. The fast portability of health data, along with the complexity of legal regulations and voluminous “Terms of Use” documents that are rarely read by users,^30^ create a reality of data that has the potential to quickly bounce to all corners of the world. In addition, there is the very real presence of hackers.^31^ Therefore, some of the data receivers have motives that have nothing to do with “public interest.” Accordingly, the fears and lack of trust we observed are likely well-founded and highlight the need for risk mitigation to harness the full promise of public health surveillance during a pandemic.

### Strengths and Limitations

This is the first global survey, to our knowledge, to investigate the opinions of physicians about utility, trust, and risks of commonly used public health digital surveillance tools. Our survey data is from a relatively large and diverse sample of verified practicing physicians. Potential limitations include cross-sectional design, limited number of respondents from developing countries, inability to control for all possible confounding variables (e.g., personal medical history, socio-political beliefs, local data privacy regulations, knowledge about digital tools), and inability to deduce causality. Further, physician perceptions may change over time if infection risk and prevalence decrease, due to vaccination and herd immunity. Our findings should be interpreted within this context. Nevertheless, they provide a useful baseline for future surveys.

### Public Health Implications

Physicians were optimistic but not equally supportive of all surveillance tools suggesting the need for further research on effectiveness. There was also variation in physician opinions by age group. This may in part reflect differences in physician knowledge about emerging technologies and/or risk-benefit analyses, which would benefit from further education. The low level of trust in technology companies to protect personal data suggests that independent entities (governed by stricter privacy laws) should be the gatekeepers of such data. Current regulations fall short of addressing the risks posed by these new technological developments. It has been said that “data moves at the speed of trust”. During public health emergencies any data collection through such newer tools should be both time-limited and scope-limited, with decisions made in a transparent way prior to the launch of surveillance activity.^22,27^ In parallel, we may need to strengthen other data privacy rules to ensure any temporary loosening during public health emergencies does not result in future misuse in normal times. We hope that insights from surveys such as this may spur public health agencies and technology innovators to work together to develop the evidence base and balance individual versus societal versus commercial needs.^24^ As aptly noted by one of the survey respondents, “we can learn from films like Spiderman and The Dark Knight – with great power comes great responsibility”.

## Data Availability

Data will be made available upon request.

## Acknowledgments

The authors thank Sermo and express gratitude to the physicians who shared their insights. We also thank Jolynn Dellinger at the Kenan Institute for Ethics for her insights.

