## Supplemental Table 1 for "Physician Perceptions of Surveillance: Wearables, Apps, and Bots for COVID-19"

**Supplemental Table 1. Survey Questions Reported in the Study**

Contact tracing apps on smart phones are being promoted by governments to monitor and curb the spread of COVID-19. Such apps use your personal identity and cell phone GPS signals to track your movements, the places you visited and who you have been in contact with for the past 2 weeks. Do you believe that the possible benefits to the public of COVID-19 contact tracing apps will outweigh the possible risks/harms?
*Yes/No/Uncertain*

Smart watches and wearables may soon be able to predict if a person has an infection before they can self-recognize it (pre-symptomatic stage) based on changes in temperature, heart rate, respiration and sleep. If 15% of the population have an infection, a device that is 90% accurate will have a 96% negative predictive value and a 60% positive predictive value. Do you believe that the possible benefits of such an infection detection alert will outweigh the possible risks/harms? *Yes/No/Uncertain*

"Fever cameras" (Infrared Temperature Detection Systems) are being used for mass screening of the public for fever (e.g. at airports, schools). Current research suggests that such systems have 85% accuracy, a 99% negative predictive value and a 15% positive predictive value for detecting true fever. Do you believe that the possible benefits of such a mass fever screening system will outweigh the possible risks/harms? *Yes/No/Uncertain*

Chatbots were used by large numbers of people during COVID-19 to screen themselves for symptoms and determine the need for further evaluation. In most countries, such chatbots do not need prior regulatory approval. *Do the possible benefits of such COVID-19 symptom screening chatbots outweigh the possible risks/harms? Yes/No/Uncertain*

Social media postings (e.g. on Facebook) are being aggregated to create "COVID-19 symptom maps" and population movement maps. Anonymized maps are being shared with public health officials and researchers. Do the possible benefits of such social media tracking outweigh the possible risks/harms? *Yes/No/Uncertain*

If COVID-19 surveillance technologies were offered, how much would you trust the following groups to protect your privacy? (Choose one)
*Technology companies; Government; My employer; My medical provider; educational universities/non-profit bodies; I would not trust anyone to protect my data privacy*

If privacy standards are loosened to fight the pandemic, do you believe that in the future this will also lead to misuse of privacy in other areas of personal life? *Yes/No/Uncertain*
