## Supplemental Table 2 for "Physician Perceptions of Surveillance: Wearables, Apps, and Bots for COVID-19"

**Supplemental Table 2. Country where the Respondent Physician Practices (N=1004).**

| Country | N | % |
| --- | --- | --- |
| United States | 340 | 33.9% |
| Italy | 116 | 11.6% |
| Spain | 83 | 8.3% |
| United Kingdom | 53 | 5.3% |
| Germany | 47 | 4.7% |
| Mexico | 44 | 4.4% |
| France | 40 | 4.0% |
| Russia | 36 | 3.6% |
| Canada | 33 | 3.3% |
| Brazil | 30 | 3.0% |
| Venezuela | 30 | 3.0% |
| India | 27 | 2.7% |
| China | 18 | 1.8% |
| Australia | 17 | 1.7% |
| Turkey | 14 | 1.4% |
| Japan | 12 | 1.2% |
| Portugal | 9 | 0.9% |
| South Korea | 7 | 0.7% |
| South Africa | 7 | 0.7% |
| Poland | 6 | 0.6% |
| Belgium | 5 | 0.5% |
| Greece | 5 | 0.5% |
| Argentina | 3 | 0.3% |
| Philippines | 2 | 0.2% |
| Switzerland | 2 | 0.2% |
| Austria | 2 | 0.2% |
| Colombia | 2 | 0.2% |
| Sweden | 2 | 0.2% |

*One respondent (0.1%) each from Malaysia, the Netherlands, Honduras, Jordan, New Zealand, Ukraine, Czech Republic, Kazakhstan, United Arab Emirates, Hungary, Saudi Arabia, and Nigeria are not shown in the Table.
