## Supplemental Table 3 for "Physician Perceptions of Surveillance: Wearables, Apps, and Bots for COVID-19"

**Supplemental Table 3. Selected Comments Supportive of Digital Surveillance Tools**

- During the 1940-41 bombing of London called The Blitz, I believe there were zero residents of London who said "I have a constitutional right to leave my lights on a night if I feel like it.”
- Generally speaking, I favor almost all methods for identification, contact tracing, and isolation.
- I believe it will become a necessity in order to flatten the curve and prevent rampant infections.
- They are helpful for healthy travelers and commuters.
- Using the phone to warn possible exposure would be helpful when you are in area outside your routine or interact with many people.
- The Future is Here
- I believe that in the future digital surveillance technology will be indispensable.
- I believe that a full-fledged record of contact and sick people with their timely isolation is the best way to localize foci. And using digital tracking technologies to do this is a good way to make this function complete.
- Digital surveillance technologies will play an increasingly important role in current and future pandemics.
- Anything that prevents deaths is fine with me. I don't worry about privacy
- There is already extensive digital surveillance and data sharing for law enforcement and mercantile purposes, so why not harness it for the public health
- It works! We have seen it work in other countries
- Using it to map areas of high activity makes sense (home address, not the location the positive test occurred).
- The Unstoppable Future. It must be mandatory, not optional
