## Supplemental Table 4 for "Physician Perceptions of Surveillance: Wearables, Apps, and Bots for COVID-19"

| **Supplemental Table 4. Selected Comments About Efficacy Gaps with Surveillance Tools** |
|---|
| • Digital surveillance is NOT ready for prime time. Too much risk for: abuse, false results causing more confusion. Would favor if the PPV and NPV were = 95%<br>• We do not know enough about the virus, and technologies are not enough reliable.<br>• Way too early. Data should be accumulated and analyzed in context of controlled trials by reputable scientists.<br>• I fear the behavior of some people, not the technologies. Need deep cultural change for compliance<br>• The bad will eventually outweigh the initial good. There MUST be an ending period established before it is employed<br>• Bad for patient and physician; Useless. IT is not medicine<br>• It's not the acquisition of the data that concerns me, but more those who collect and analyze .<br>• None of them can perform better than an accurate history of symptoms, and the persons ability to refrain from locally spreading their germs<br>• Better leadership and reliance on scientific medicine is far more important than unproven and invasive technologies. |
