## Supplemental Table 5 for "Physician Perceptions of Surveillance: Wearables, Apps, and Bots for COVID-19"

| **Supplemental Table 5. Selected Comments about Ethics, Privacy and Harms of Surveillance** |
| --- |
| • Afraid of where we are going…I was afraid BEFORE Covid, even more so now.<br>• I think digital surveillance is a slippery slope. First COVID, then what is next?<br>• Big Brother is salivating. Re-read 1984. Read Hariri´s 21 lessons for the 21st century<br>• The winners will be private investors...<br>• Digitalize schools and universities, not Daily Life<br>• Only totalitarian governments are able to control and surveil this kind pandemics.<br>• A LOT OF POWER TO THE STATE, AND THAT'S NOT GOOD FOR ANYONE<br>• Like the atomic bomb, any great idea can have unforeseen consequences<br>• Tech companies have no conscience, and compensate for that by overemphasizing support of liberal programs.. The rights and wellbeing of the average citizen have no place in their thinking. ..will never get our privacy back once it has been given away.<br>• Too much potential for corruption and misuse. Violates essential freedoms<br>• This country is based on personal freedoms. This is a significant invasion of privacy. Will need Socialism for this to work. I am against that.<br>• Creepy. Extreme slippery slope. We've already given up so very many of our privacy rights in our society.<br>• Setting a precedent for abuse. Don't even give an inch because they will take a mile;<br>• I am very concerned about privacy issues and that any "temporary" relaxation of standards will become permanent.<br>• Digital surveillance technologies in the hands of an autocratic government are a slippery slope to the loss of liberty.<br>• These tools will facilitate discrimination and other rights abuse against racial minorities and people living in poverty<br>• Pandora's box<br>• Lots of potential for corruption. Criminals and corrupt politicians will use this to track their unsuspecting victims<br>• We can use incredible technology for the benefit of mankind, the problem we have is we keep handing this power to whoever can afford it, not those who deserve it. |
